# Proxalutamide Improves Inflammatory, Immunologic, and Thrombogenic Markers in Mild-to-Moderate COVID-19 Males and Females: an Exploratory Analysis of a Randomized, Double-Blinded, Placebo-Controlled Trial Early Antiandrogen Therapy (EAT) with Proxalutamide (The EAT-Proxa Biochemical AndroCoV-Trial)

**DOI:** 10.1101/2021.07.24.21261047

**Authors:** Flávio Adsuara Cadegiani, Andy Goren, Carlos Gustavo Wambier, Ricardo Ariel Zimerman

## Abstract

**Background:** The androgen theory on COVID-19 is based on the fact that males, in particular when affected by androgenetic alopecia, and females with hyperandrogenic states are more severely affected by COVID-19, while chronic users of antiandrogens experiment lower rates of COVID-19 complications. The theory finds plausibility on the androgen-mediated transmembrane protease serine 2 (TMPRSS-2), a key protein for SARS-CoV-2 cell entry. We demonstrated reduction of hospitalization rate using a potent non-steroidal antiandrogen (NSAA), proxalutamide, in both females and males COVID-19 outpatients. In this joint exploratory analysis, we aimed to demonstrate whether the efficacy of proxalutamide on mild-to-moderate COVID-19 could be justified by improvements in inflammatory, immunologic, and thrombogenic responses.

**Methods:** This is a joint *post-hoc* analysis of two double-blind, placebo-controlled two-arm randomized clinical trials (RCTs) on proxalutamide 200mg/day for seven days for female and male COVID-19 outpatients, respectively, compared to standard of care (SOC), of hematocrit, neutrophils lymphocytes, eosinophils, platelets, neutrophil-to-lymphocyte (N/L) ratio, ferritin, fibrinogen, D-dimer, ultrasensitive C-reactive protein (usCRP) lactate 1-hour erythrocyte sedimentation rate (1hESR), total testosterone, estradiol, sex hormone binding globulin (SHBG), oxygen saturation and heart rate measured on days 0, 1 and 7.

**Results:** A total of 445 subjects were enrolled (268 males and 177 females) between October 21^th^ 2020 and February 28^th^ 2021, with similar baseline characteristics. Neutrophils were lower in proxalutamide group in Day 1 (p = 0.005) and Day 7 (p < 0.0001). Lymphocytes were higher in the proxalutamide group in Day 7 (p = 0.0001). Eosinophils were higher in the proxalutamide arm in Day 1 (p = 0.04) and Day 7 (p < 0.00010. In Day 7, platelets were higher in proxalutamide arm (p = 0.03). Ferritin levels were lower in proxalutamide arm in Day 7 (p = 0.03) Fibrinogen levels were lower in proxalutamide group in Days 1 and 7 (p < 0.0001 for both days). D-dimer levels were lower in proxalutamide group in Days 1 and 7 (p < 0.0001 for both days). UsCRP levels were reduced in proxalutamide group in Day 7 (p < 0.0001). 1hESR) was reduced in proxalutamide arm in Day 1 (p = 0.0009) and Day 7 (p < 0.0001). In males, testosterone levels were higher in proxalutamide group in Day 1 (p = 0.048) and Day 7 (p = 0.0001). In females, testosterone levels were higher in proxalutamide group in Day 7 (p = 0.018), and estradiol levels were higher in proxalutamide arm in Day 1 (p = 0.044). Oxygen saturation was higher in proxalutamide in Day 1 (p = 0.0006) and Day 7 (p < 0.0001).

**Conclusions:** The substantial improvements observed in immunologic, inflammatory, thrombotic and oxygen markers with proxalutamide may support the reduction of hospitalization rate observed in both females and males with COVID-19 using proxalutamide, compared to standard of care.

## Introduction

COVID-19 pandemic has affected males disproportionally in terms of severity and mortality, even after adjustments for age, body mass index (BMI), and comorbidities.^1^ Among males, those affected by androgenetic alopecia (AGA), a phenotypical expression of androgen hyperactivity, demonstrated to be more severely affected, irrespective of other risk factors.^2-5^ Pre-pubertal children, unexposed to androgens, are less affected, while babies under one year of age have relatively higher risk of COVID-19 as compared to older pre-pubertal children.^6^ This may be related to the existence of a physiological occurrence of mini-puberty at this age, when the gonadotropic axis is unblocked.^6^ Finally, women with hyperandrogenic phenotypes, including polycystic ovary syndrome (PCOS), were shown to be at higher risk for severe COVID-19.^7,8^ Hence, overwhelming epidemiological and observational data has pointed to androgens as being key for SARS-CoV-2 infection severity.

In addition to the strong epidemiology associating androgenic phenotypes and COVID-19 severity, emerging data demonstrate that targeting androgens have become a promising therapeutical tool for COVID-19 based on chronic use of antiandrogens. Older males affected by severe, metastatic, castration-resistant prostate cancer, expected to be more affected by COVID-19, were actually relatively protected for COVID-19, when under androgen deprivation therapy (ADT), despite the intense sarcopenia and other complications related to long-term ADT that could be additional risk factors for COVID-19.^9-11^ Conversely, younger males taking dihydrotestosterone (DHT) derivate anabolic androgenic steroids (AAS) may be at higher risk for severe COVID-19.^12^ Collectively, these findings suggest that exposure to androgens is likely an independent risk factor for COVID-19 severity.

From the molecular and virological perspectives, SARS-CoV-2 cell entry depends on two major proteins, known as angiotensin-converting enzyme 2 (ACE-2) and transmembrane protease serine 2 (TMPRSS-2).^13^ Dysfunctions in the renin-angiotensin-aldosterone system (RAAS), with consequent inadequate ACE-2 expression, may justify why metabolic disorders, including type 2 diabetes mellitus (T2DM), hypertension, and obesity, are major risk factors for sever COVID-19, are at higher risk for severe COVID-19.^14^ However, dysfunctions in the ACE-2 expression may not fully justify all risk factors identified for severe COVID-19.^14^

TMPRSS-2 promotes cleavage of SARS-CoV-2 spike protein and is essential for subsequent viral cell entry. The explanation for the observations that led to the hypothesis of androgen activity as being a predictor of COVID-19 severity^2-12^ is based on the fact that the only endogenous known modulators of TMPRSS2 synthesis are the androgens.^13,14^ Indeed, anti-androgenic approaches demonstrated preliminary evidence of protection for COVID-19, both in chronic users and when specifically administered for the disease.^15-17^ Stronger AR antagonists tend to perform better against SARS-CoV-2, suggesting dose-dependent effect and thus biological activity.^14,17^

Non-steroidal anti-androgens (NSAAs) are a class of strong-acting androgen receptor (AR) antagonists and/or suppressors of AR expression, used as ADT for castration-resistant prostate cancer, male-to-female transgender transition, hirsutism, precocious puberty and priapism.^18-20^ First generation NSAAs include flutamide and bicalutamide. Second generation have advantages over first generation in terms of potency, efficacy, and liver safety profile, and include enzalutamide, apalutamide, darolutamide and, more recently, proxalutamide.

We hypothesized that using a strong NSAA would provide protection against the likely androgen-mediated SARS-CoV-2 infection. Proxalutamide, a potent NSAA, has demonstrated to be up to ten times more potent to antagonize AR activity then older agents,^21-23^ and is the sole NSAA shown to suppress AR expression. Proxalutamide has also demonstrated to block angiotensin-converting enzyme 2 (ACE-2) directly, and to mitigate inducible Nitric Oxide Synthase (iNOS) and Tumoral Necrosis Factor alpha (TNF-alpha) expression,^21,22^ determining a potential potent anti-inflammatory effect.

As a consequence, we have conducted two double-blinded, placebo-controlled randomized clinical trials (RCTs) to test the efficacy of proxalutamide compared to placebo in mild-to-moderate COVID-19 outpatients – one in females and one in males. In accordance with our hypothesis, in both studies, substantial reductions in hospitalization rates were observed,^24,25^ supported by the previously reported antiviral and protective radiological effects of proxalutamide.^26,27^

We aimed to evaluate whether the improvements in clinical outcomes observed with the use of proxalutamide is accompanied by biochemical improvements in terms of inflammatory and thromboembolic parameters, since these are two cornerstones of the SARS-CoV-2 pathophysiology, demonstrated to be important surrogate markers of COVID-19 infections playing as independent prognostic factors. To address this question, we performed a joint exploratory analysis of the biochemical responses of the RCTs on proxalutamide in COVID-19 male and female outpatients. ^24,25^

## Materials and methods

Subjects with confirmed COVID-19 through a positive rtPCR-SARS-CoV-2 test for the past seven days were recruited to participate in the present randomized, double-blinded, placebo-controlled clinical trial to test the efficacy of proxalutamide, a second generation NSAA, versus standard of care (SOC) solely. Inclusion criteria included age above 18 years old, absence of contraindication to any of the drugs used in the study, oxygen saturation above 92% and confirmed non-pregnancy in case of females. Exclusion criteria included breastfeeding, signs of COVID-19 complications, including reduction of oxygen saturation above 4% or oxygen saturation below 92%, and use of specific alleged ‘treatments’ for COVID-19 for more than 48 hours, including hydroxychloroquine, ivermectin or nitazoxanide.

After the written consent form was given, subjects were designated to either active (proxalutamide) arm or placebo arm. Baseline characteristics including gender, age, BMI, comorbidities and use of medications were assessed. This RCT has been approved by the national ethics committee (approval numbers 4.173.074 and 4.513.428, for males and females, respectively). This RCT was registered in clinicaltrials.gov: numbers NCT04853134 and NCT04446429, for females and males, respectively. The study was conducted at two study sites (Corpometria Institute, Brasilia, Brazil, and Laboratorio Exame, DASA, Brasilia, Brazil) by the principal investigator (PI), and coordinated by the study director.

Males and females were randomized in 1:1 ratio between proxalutamide and placebo, in blocks of six and 25, respectively. Whenever subjects were not using specific drugs for COVID-19, standard of care (SOC), tconsisted of nitazoxanide 500mg twice daily after meals for six days plus azithromycin 500mg daily for five days, and symptomatic drugs, were given. Subjects in the active arm were given proxalutamide 200mg daily for up to seven days (all females received for seven days), and the placebo arm received an identical placebo pill daily for seven days.

The present analysis is a *post-hoc* report of the biochemical parameters and vital signs of the RCT in both males and females. Biochemical parameters recorded include hematocrit (%), neutrophils (* 1000/mm^3^), lymphocytes (* 1000/mm^3^), eosinophils (* 1000/mm^3^), and platelets (* 1000/mm3) (flow cytometry with fluorescent protein - XN10-Sysmex), ferritin (ng/mL) (chemiluminescence - CLIA), fibrinogen (mg/dL) (Clauss assay), D-dimer (ng/mL) (immunologic assay), ultrasensitive C-reactive protein (usCRP, mg/L) (latex-intensified immunoturbidimetry), lactate (mmol/L) (enzymatic assay), erythrocyte sedimentation rate (ESR, mm/1h) (capillary photometry), total testosterone (ng/dL) (electric chemiluminescence - ECLIA), estradiol (pg/mL) (ECLIA), and sex hormone binding globulin (SHBG) (nmol/L) (ECLIA), on Day 0, Day 1, and Day 7. Neutrophil-to-lymphocyte ratio was then calculated for Day 0, Day 1, and Day 7. A new RT-PCR-SARS-CoV-2 kit testing (Automatized Platform, Roche USA, Indianapolis, IN) following the COBAS SARS-CoV-2 RT-PCR kit test protocol was collected in Day 7 by some of the subjects. Baseline 25(OH) vitamin D (ng/mL) (ECLIA), homocysteine (umol/L) (ECLIA), and glycated hemoglobin (HbA1c) (%) (turbidimetric immunoassay) were collected. Oxygen saturation (%) and heart rate (bpm) were measured in Day 0, Day 1, and Day 7. Analyses were performed for overall and stratified for gender.

### Statistical analysis

Baseline characteristics, medical history, comorbidities, use of medications were checked for similarity between active and placebo arms. Biochemical and vital sign data was checked for normality. Parametric parameters were presented as means and standard deviations (SD) and non-parametric parameters were presented as medians and interquartile ranges (IQR). A non-parametric statistical tool (Mann-Whitney U test) was employed to determine statistical significance for all parameters, which was set at p < .05. The XLSTAT version 2020.3.1.1008 (Addinsoft, Inc. New York, NY) was employed for the statistical analysis. All missing data are Missing Completely at Random (MCAR), except for six male patients that dropped out, although their outcomes were known and counted for intention-to-treat (ITT) analysis for hospitalization rate elsewhere.^24,25^ The amount of missing data was provided for each parameter at each time. However, treatment effect for hospitalization rate was calculated adjusted for missing data, without significant differences. Therefore, missing data unlikely biased the analysis of parameters evaluated in this study.

## Results

A total of 445 subjects were enrolled, including 268 males and 177 females that were randomized between October 21^th^ 2020 and February 28^th^ 2021. Of the 268males, 134 were designated for the active arm and 134 for the placebo arm. Of the 177 females, 75 were designated for the active arm and 236 for the placebo arm. Overall, 209 patients were randomized to proxalutamide and 68 were randomized to placebo.

Baseline characteristics distributions between groups were as previous published. Details can be found in Table 1. Patients did not show statistically differences in demographic or clinical characteristics. Results of the biochemical parameters are described in Table 2 and illustrated in Figure 2.

**Table 1:** Baseline characteristics.

| Characteristics | All<br>(n=445)<br>Female (n=177)<br>Male (n=268) | Proxalutamide<br>(n = 209)<br>Female (n=75)<br>Male (n=134) | Placebo<br>(n =236)<br>Female (n=102)<br>Male (n=134) |
| --- | --- | --- | --- |
| Age (years), median (IQR) | 45 (17)<br>Female – 44 (17)<br>Male – 45 (17) | 45 (19)<br>Female – 45 (17)<br>Male – 45 (19) | 45 (15)<br>Female - 43 (16)<br>Male – 46 (15) |
| Race |  |  |  |
| Mixed ethnicity* | 100% (445) | 100% (209) | 100% (236) |
| No. of coexisting conditions —<br>% (Number) |  |  |  |
| None | 58% (259)<br>Female - 57% (101)<br>Male – 59% (158) | 54% (112)<br>Female - 51% (38)<br>Male – 55% (74) | 62% (147)<br>Female - 62% (63)<br>Male – 63% (84) |
| One | 22% (98)<br>Female - 28% (49)<br>Male – 18% (49) | 24% (51)<br>Female - 27% (22)<br>Male – 22% (29) | 20% (47)<br>Female - 27% (27)<br>Male – 15% (20) |
| Two or more | 18% (82)<br>Female - 15% (27)<br>Male – 21% (55) | 22% (46)<br>Female - 20% (15)<br>Male – 23% (31) | 15% (36)<br>Female - 12% (12)<br>Male – 18% (24) |
| Coexisting conditions —<br>% (Number) |  |  |  |
| Type 2 diabetes | 8% (36)<br>Female - 9% (15)<br>Male – 8% (21) | 9% (18)<br>Female - 9% (7)<br>Male – 8% (11) | 8% (18)<br>Female - 8% (8)<br>Male – 7% (10) |
| Hypertension | 21% (94)<br>Female - 22% (39)<br>Male – 21% (55) | 24% (51)<br>Female - 24% (18)<br>Male – 25% (33) | 18% (43)<br>Female - 21% (21)<br>Male – 16% (22) |
| COPD | 0% (2)<br>Female - 1% (1)<br>Male – 0% (1) | 1% (2)<br>Female - 1% (1)<br>Male – 1% (1) | 0<br>Female – 0<br>Male – 0 |
| BMI > 30 kg/m<sup>2</sup> | 17% (75)<br>Female - 18% (32)<br>Male – 16% (43) | 17% (36)<br>Female - 19% (14)<br>Male – 16% (22) | 17% (39)<br>Female - 18% (18)<br>Male – 16% (21) |

BMI = body mass index P > 0.05 for all characteristics.

**Table 2.**
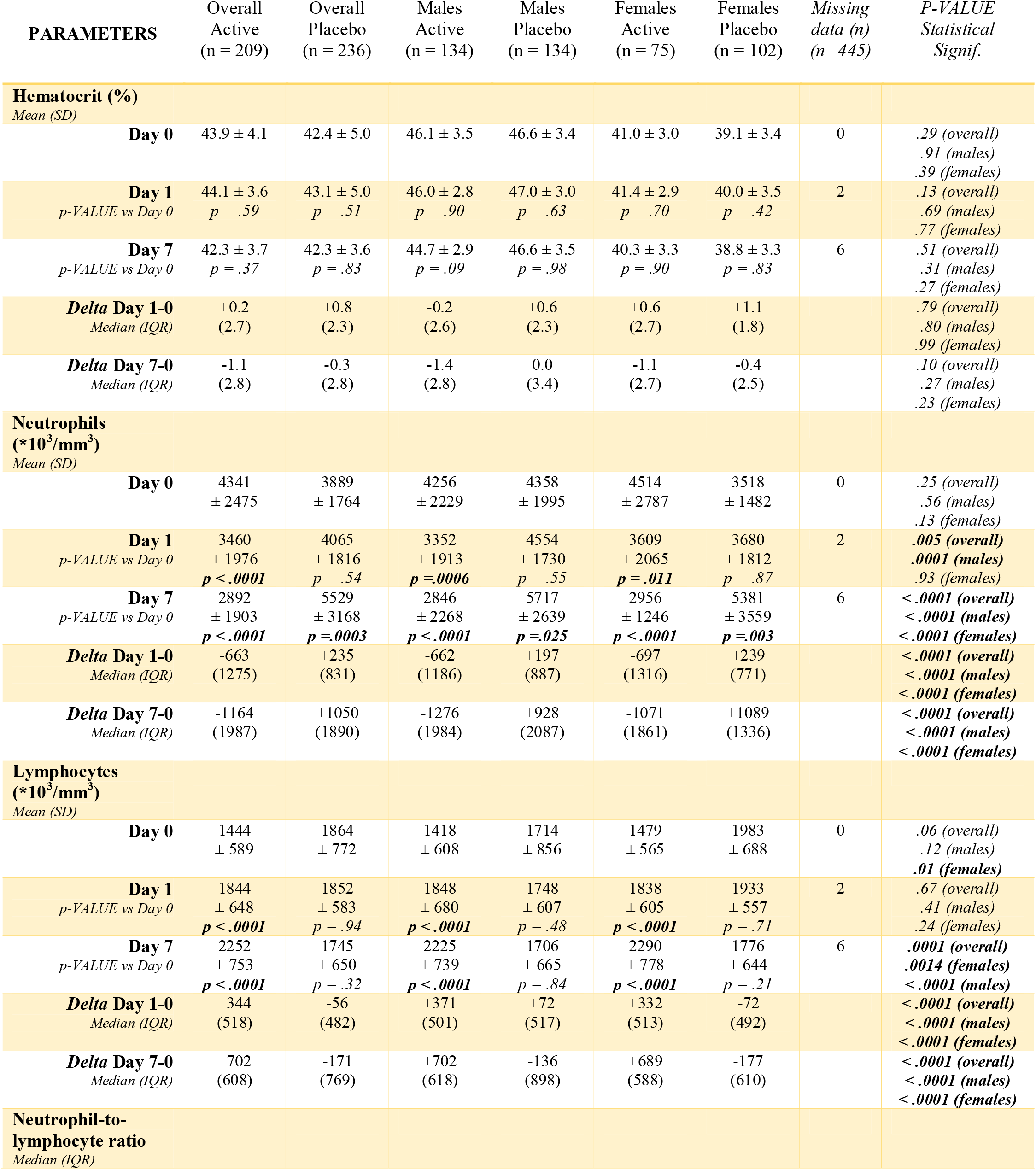
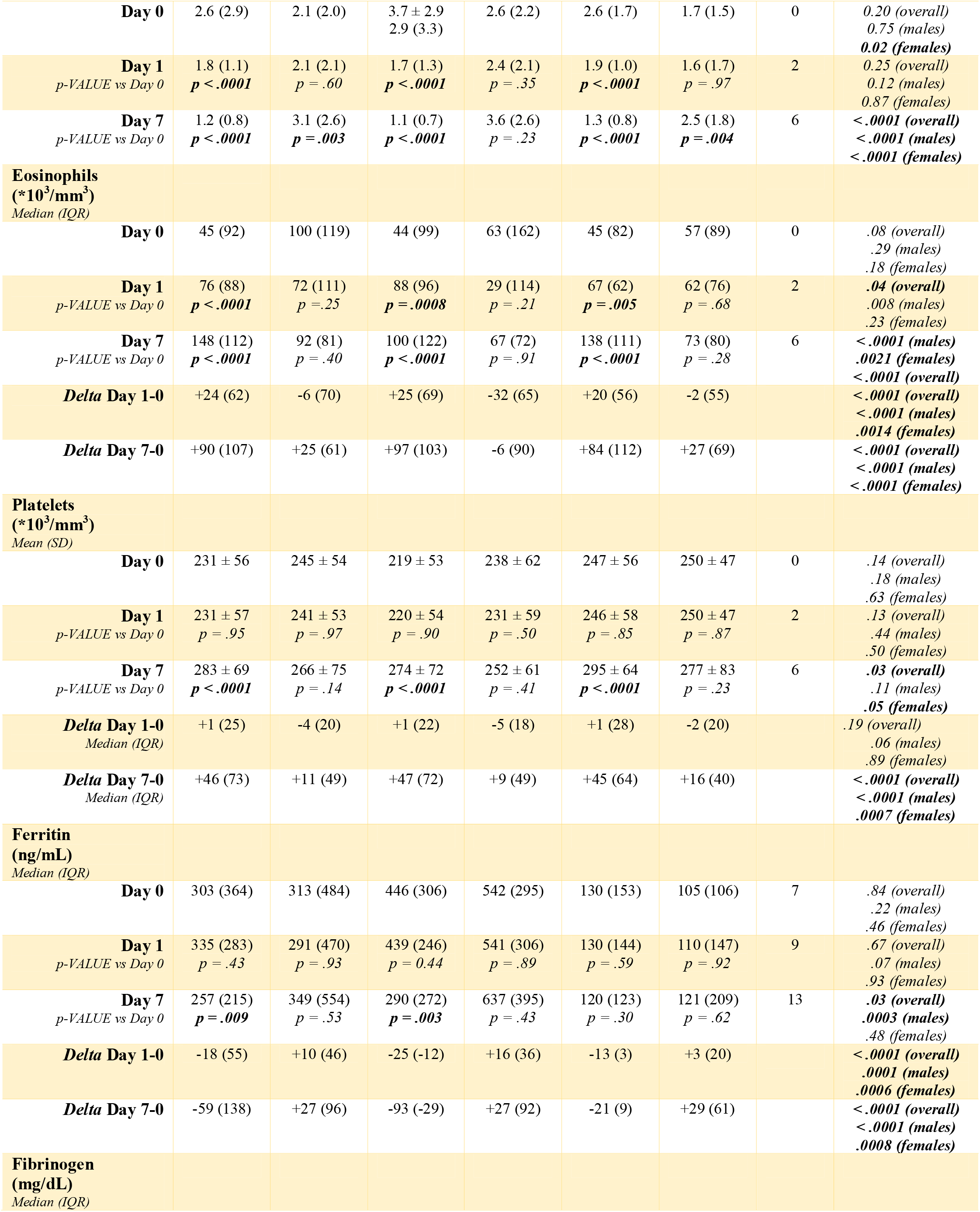
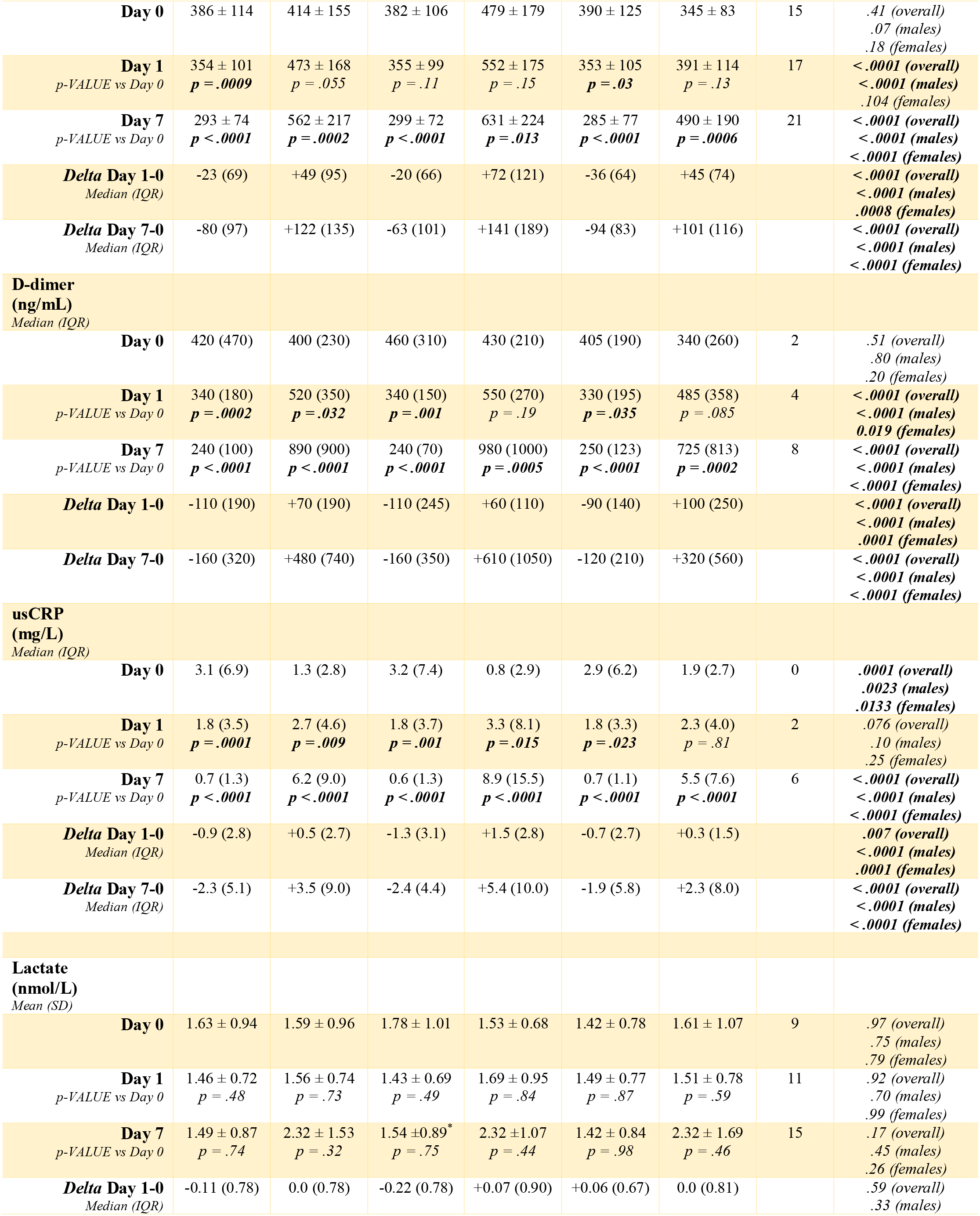
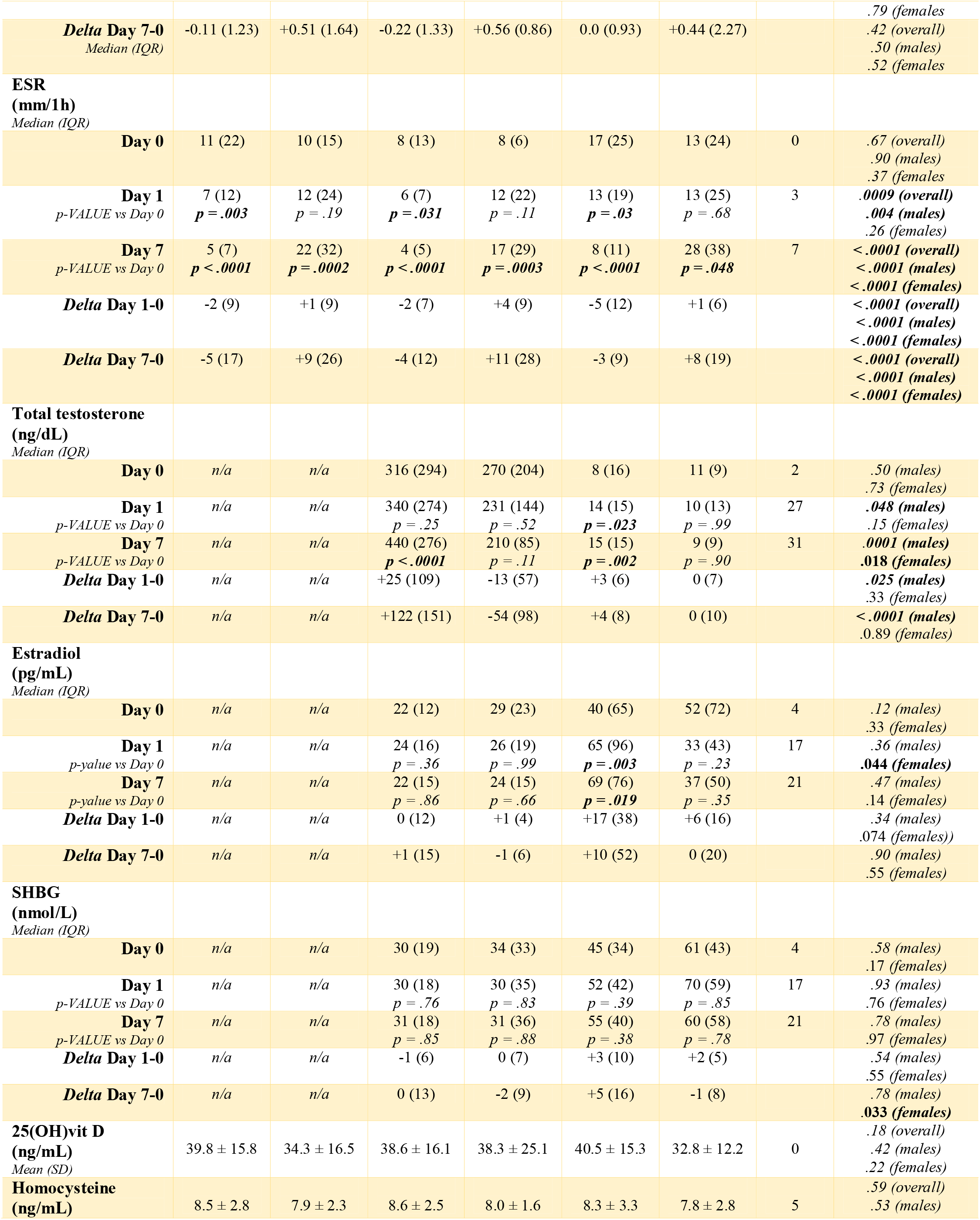
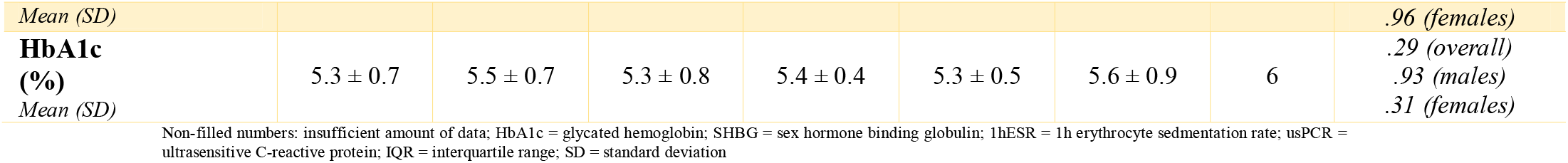
Baseline and study outcomes stratified by sex, day of follow-up and variation between Day 1 and Day 0 and between Day 7 and Day 0.

**Table 3.** Vital signs stratified by sex, day of follow-up and variation between Day 1 and Day 0 and between Day 7 and Day 0.

| PARAMETERS | Overall<br>Active<br>(n = 209) | Overall<br>Placebo<br>(n = 236) | Males<br>Active<br>(n = 134) | Males<br>Placebo<br>(n = 134) | Females<br>Active<br>(n = 75) | Females<br>Placebo<br>(n = 102) | Missing<br>data (n)<br>(n=445) | P-VALUE<br>Statistical<br>Signif. |
| --- | --- | --- | --- | --- | --- | --- | --- | --- |
| **Oxygen saturation (%)** |  |  |  |  |  |  |  |  |
| Mean (SD) |  |  |  |  |  |  |  |  |
| **Day 0** | 96.3 ± 2.0 | 96.8 ± 1.5 | 95.9 ± 2.2 | 95.8 ± 1.5 | 96.7 ± 1.7 | 97.3 ± 1.6 | 0 | .93 (overall)<br>.12 (males)<br>.068 (females) |
| **Day 1**<br>*p-VALUE vs Day 0* | 97.2 ± 1.5<br>*P < .0001* | 96.3 ± 1.9<br>*P = .12* | 97.0 ± 1.6<br>*P = .0012* | 96.2 ± 2.2<br>*P = .86* | 97.5 ± 1.3<br>*P = .0078* | 96.4 ± 1.4<br>*P = 0.01* | 38 | .0006 (overall)<br>.082 (males)<br>.0007 (females) |
| **Day 7**<br>*p-VALUE vs Day 0* | 97.9 ± 1.4<br>*P < .0001* | 96.7 ± 2.0<br>*P = .85* | 97.7 ± 1.4<br>*P < .0001* | 96.5 ± 2.2<br>*P = .30* | 98.0 ± 1.4<br>*P < .0001* | 96.8 ± 1.8<br>*P = 0.27* | 6 | .0001 (overall)<br>.008 (males)<br>.0001 (females) |
| **Delta Day 1-0**<br>*Median (IQR)* | +1 (2) | -1 (3) | +1 (2) | 0 (2) | +1 (1.5) | -1 (3) |  | < .0001 (males)<br>< .0001 (females)<br>< .0001 (overall) |
| **Delta Day 7-0**<br>*Median (IQR)* | +1 (3) | 0 (2) | +2 (3) | +0.5 (3) | +1 (2) | 0 (2) |  | < .0001 (males)<br>< .0001 (females)<br>< .0001 (overall) |
| **Heart rate (bpm)** |  |  |  |  |  |  |  |  |
| Mean (SD) |  |  |  |  |  |  |  |  |
| **Day 0** | 78.4 ± 12.2 | 75.6 ± 11.2 | 76.8 ± 11.5 | 76.0 ± 13.5 | 80.7 ± 12.9 | 75.3 ± 9.3 | 0 | .16 (overall)<br>.43 (males)<br>.14 (females) |
| **Day 1**<br>*p-VALUE vs Day 0* | 80.7 ± 12.8<br>*P = .17* | 82.6 ± 13.1<br>*P = .002* | 78.8 ± 11.6<br>*P = .38* | 84.5 ± 14.8<br>*P = .018* | 83.5 ± 14.0<br>*P = .31* | 81.3 ± 11.7<br>*P = .044* | 44 | .35 (overall)<br>.08 (males)<br>.57 (females) |
| **Day 7**<br>*p-VALUE vs Day 0* | 78.4 ± 12.8<br>*P = .39* | 79.2 ± 13.1<br>*P = .10* | 77.2 ± 11.4<br>*P = .94* | 78.3 ± 13.1<br>*P = .45* | 80.3 ± 14.5<br>*P = .22* | 87.0 ± 13.6<br>*P = .10* | 6 | .73 (overall)<br>.92 (males)<br>.38 (females) |
| **Delta Day 1-0**<br>*Median (IQR)* | +3 (16) | +6 (10) | +3 (16) | +7 (11) | +2 (15) | +5 (11) |  | .0016 (overall)<br>.0035 (males)<br>.07 (females) |
| **Delta Day 7-0**<br>*Median (IQR)* | -1 (19) | +2 (10) | -1 (17) | +2 (5) | -1 (20) | +2 (17) |  | .08 (overall)<br>.52 (males)<br>.07 (females) |

Hematocrit (Ht) did not demostrate differences between proxalutamide and placebo in Day 0, Day 1, and Day 7, as well as in the differences between Days 1 and 0 and between Days 7 and 0, for females, males, and overall patients.

Basal absolute neutrophil counts (ANC) were similar between groups at baseline. Overall proxalutamide patients had significant lower ANC than the placebo group in Day 1 (p = 0.005) and Day 7 (p < 0.0001). Correspondingly,, ales on the proxalutamide arm had significant lower ANC compared to placebo males in Day 1 (p = 0.0001) and Day 7 (p < 0.0001). Females of the proxalutamide arm had similar levels than placebo females in Day 1, but significantly lower levels in Day 7 (p < 0.0001). Absolute changes of ANC between Day 1 and Day 0 and between Day 7 and Day 0 were negative for overall and sex subgroups in the proxalutamide arm, and positive for overall and sex subgroups in the placebo arm, with significant differences in the changes between proxalutamide and placebo arm at all times for all groups (p < 0.0001 for all). In the proxalutamide group, significant reduction of ANC was observed in Day 1 compared to Day 0 in overall (p < 0.0001), females (p = 0.011), and males (p = 0.0006), as well as Day 7 compared to Day 0 (p < 0.0001 for overall, males, and females). In the placebo arm, significant increase of ANC was observed between Day 7 and Day 0 in overall (p = 0.0003), females (p = 0.003) and males (p = 0.025).

Baseline total lymphocytes count (TLC) were similar in males but significantly reduced in females of the proxalutamide group compared to the placebo group (p = 0.01) and marginally non-significantly reduced in overall patients in the proxalutamide arm (p = 0.06). In Day 1, TLC was similar between groups. In Day 7, TLC was significantly higher in the proxalutamide group than in the placebo group for overall (p = 0.0001), females (p = 0.0014) and males (p < 0.0001). Changes between Day 1 and Day 0 and between Day 7 and Day 0 were significantly higher towards increase in TLC in proxalutamide group (p < 0.0001 for overall, females, and males, for Days 1-0 and Days 7-0). TLC increase between Days 1 and 0 and between Day 7 and Day 0 were significant in the proxalutamide arm (p < 0.0001 for overall, females, and males, for both Days 1-0 and Days 7-0), while non-significant changes were observed in the placebo arm.

The neutrophil-to-lymphocyte (N/L) ratio was similar between groups in overall and sex subgroups at baseline and in Day 1. In Day 7, N/L ratio was significantly lower in proxalutamide group than in placebo group (p < 0.0001 for overall and female and male subgroups). N/L ratio was significantly reduced in Days 1 and 7 compared to Day 0 in proxalutamide arm (p < 0.0001 for both Days 1 and 7 for overall, females, and males), while was significantly higher in placebo arm between Day 7 and Day 0 in overall (p = 0.003) and in males (p < 0.0001).

Eosinophils were similar between proxalutamide and placebo groups at baseline. In Day 1, eosinophils were significantly higher in the proxalutamide arm compared to the placebo arm in overall (p = 0.04) and males (p = 0.008). In Day 7, eosinophils were significantly higher in proxalutamide compared to placebo (p < 0.0001 for overall, females, and males). Changes in eosinophils between Days 1 and 0 and between Days 7 and 0 were significantly higher towards increase in proxalutamide arm than in placebo arm (p < 0.0001 for overall and males in Day 1 and Day 7, and females in Day 7, and p = 0.001 in Day 1 for females). Eosinophils reduced significantly in proxalutamide arm between Day 1 and Day 0 in overall (p < 0.0001), females (p = 0.005), and males (p = 0.0008), as well as between Day 7 and Day 0 (p < 0.0001 for overall, females, and males). Conversely, eosinophils did not change significantly in the placebo arm between Days 1 and 0 and between Days 7 and 0, in overall, females, and males.

Platelets were similar between proxalutamide and placebo arms at baseline and in Day 1 in overall, females, and males. In Day 7, platelets were significantly increased in proxalutamide arm compared to placebo arm in overall (p = 0.03) and females (p = 0.05), and non-significantly higher in males (p = 0.11). Increases in platelets were significantly more prominent in proxalutamide group compared to placebo group in overall (p < 0.0001), females (p = 0.0007), and males (p < 0.0001). Increase in platelets was significant in the proxalutamide arm in Day 7 compared to Day 0 (p < 0.0001 for overall, females, and males).

Ferritin levels were similar between proxalutamide and placebo arm in Day 0 and Day 1, and lower in proxalutamide arm in Day 7 in overall subjects (p = 0.03) and males (p = 0.0003). Ferritin had lower increases between Days 1 and 0 and between Days 7 and 0 in proxalutamide arm compared to placebo arm, in overall population (p < 0.0001 for both Days 1-0 and 7-0), females (p = 0.0006 and 0.0008 for Days 1-0 and 7-0, respectively), and males (p = 0.0001 and < 0.0001 for Days 1-0 and 7-0, respectively). Significant reduction of ferritin levels were observed between Day 7 and Day 0 in overall population (p = 0.009) and males (p = 0.003).

Baseline fibrinogen levels were statistically similar between groups. In Days 1 and 7, fibrinogen levels were significant lower in the proxalutamide group in overall population (p < 0.0001 for both days) and males (p < 0.0001 for both days), while was significantly lower in females in Day 7 (p < 0.0001). Changes between Days 1 and 0 and between Days 7 and 0 were significantly higher towards reduction of levels in proxalutamide group in overall population, females, and males [p < 0.0001 for all, except for females in Days 1-0 (p = 0.0008)]. Significant reductions of fibrinogen levels were observed in proxalutamide group between Days 1 and 0 and between Days 7 and 0 in overall subjects (p = 0.0009 and p < 0.0001, respectively), females (p = 0.03 and p < 0.0001, respectively), and between Days 7 and 0 (p < 0.0001). Significant increase of fibrinogen levels was observed between Days 7 and 0 in overall subjects (p = 0.0002), females (p = 0.0006), and males (p = 0.013).

D-dimer levels were similar between groups at baseline. In Days 1 and 7, D-dimer was significantly lower in proxalutamide group than in placebo group in overall (p < 0.0001 for both days), females (p = 0.019 for Day 1 and p < 0.0001 for Day 7), and males (p < 0.0001 for both days). Changes towards reduction of D-dimer levels were significantly more prominent in proxalutamide arm (p < 0.0001 for overall subjects and males, and for Days 7-0 change in females, and p = 0.0001 for females between Days 1 and 0). D-dimer reduced significantly in proxalutamide group between Days 1 and 0 and between Days 7 and 0 in overall population (p = 0.0002 and p < 0.0001, respectively), females (p = 0.035 and p < 0.0001, respectively), and males (p = 0.001 and p < 0.0001, respectively). D-dimer increased significantly between Days 1 and 0 in overall subjects (p = 0.032), and between Days 7 and 0 in overall subjects (p < 0.0001), females (p = 0.0002), and males (p = 0.0005).

Ultrasensitive C-reactive protein (usCRP) levels were significantly reduced in the placebo arm compared to proxalutamide group. In Day 7, levels were significantly lower in proxalutamide compared to placebo arm (p < 0.0001 for overall, females, and males). Changes between Days 1 and 0 and between Days 7 and 0 were significantly more substantial in proxalutamide arm in overall subject (p < 0.0001 for both days), females (p =0.0001 in Days 1-0 and p < 0.0001 in Days 7-0), and males (p = 0.0001 in Days 1-0 and p < 0.0001 in Days 7-0). UsCRP levels descreased significantly in the first and seventh day of treatment in proxalutamide group in overall population (p = 0.0001 and p < 0.0001, respectively), females (p = 0.023 and p < 0.0001, respectively), and males (p = 0.001 and p < 0.0001, respectively).

Lactate levels were similar between proxalutamide and placebo arms at baseline, Day 1, Day 7, changes between Days 1 and 0 and between Days 7 and 0. Lactate did not change significantly between Days 1 and 0 and between Days 7 and 0 in proxalutamide and placebo groups.

Erythrocyte sedimentation rate at hour 1 (1h-ESR) was similar between groups at baseline. In Day 1, levels were significantly reduced in proxalutamide arm compared to placebo arm in overall subjects (p = 0.0009) and males (p = 0.004). In Day 7, 1h-ESR levels were significantly lower in proxalutamide group (p < 0.0001 for overall population, females, and males). Changes in 1h-ESR levels between Days 1 and 0 and Days 7 and 0 were significantly higher towards reduction of levels in proxalutamide group (p < 0.0001 for both days for overall population, females, and males). 1h-ESR significantly reduced between Days 1 and 0 and between Days 7 and 0 in proxalutamide group in overall population (p = 0.003 and p < 0.0001, respectively), females (p = 0.03 and p < 0.0001, respectively), and males (p = 0.031 and p < 0.0001, respectively).

The analysis of testosterone, estradiol, and SHBG was performed in a sex-specific manner due to the inherent biological differences between sexes. In males, testosterone levels were similar between groups at baseline, while was higher in proxalutamide group than in placebo group in Day 1 (p = 0.048) and Day 7 (p = 0.0001). Changes in testosterone were significantly different between proxalutamide and placebo arm in Day 1 (p = 0.025) and Day 7 (p < 0.0001). Testosterone increased significantly in Day 7 compared to Day 0 in proxalutamide group (p < 0.0001). In females, basal and Day 1 testosterone was similar between groups, while testosterone levels were significantly higher in the proxalutamide group compared to placebo group in Day 7 (p = 0.018). Testosterone changes were similar between groups.

In males, estradiol levels were similar between groups at all times. In females, baseline and Day 7 estradiol levels were similar between groups, while levels were significantly higher in proxalutamide arm compared to placebo arm in Day 1 (p = 0.044). Changes in estradiol levels were similar between groups. Sex hormone binding globulin (SHBG) was similar in both sexes at all times. Baseline 25(OH) vitamin D, homocysteine and glycated hemoglobin (HbA1c) were similar between groups.

Oxygen saturation was similar between groups. In Day 1, oxygen saturation was significantly higher in proxalutamide group compared to placebo group in overall (p = 0.0006), in females (p = 0.0007), and non-significantly higher in males (p = 0.082). In Day 7, levels were significantly higher in proxalutamide arm compared to placebo arm (p < 0.0001 in overall and in females, and p = 0.008 in males). Changes in oxygen saturation between Day 1 and Day 0 and between Day 7 and Day 0 were significantly higher in all populations at all times (p < 0.0001 for all).

**Figure 1.**
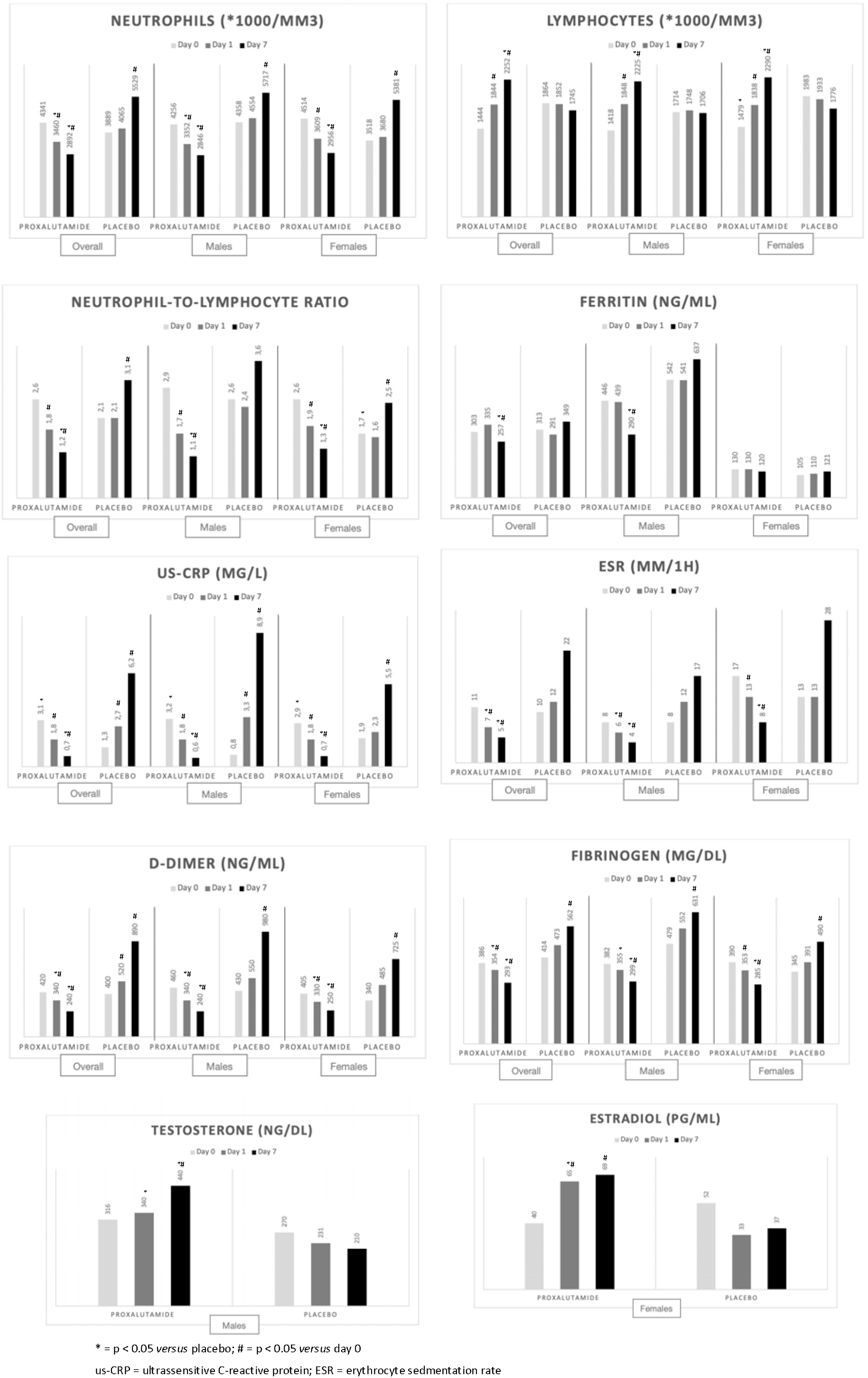
Outcomes.

## Discussion

COVID-19, caused by SARS-CoV-2, is a multisystemic disease with variable disease course, since mild clinical manifestations until need of oxygen use, mechanical ventilation, and death, besides the persistant or new-onset long-term symptoms (‘long-haulers’). In terms of prognosis, uncontrolled viral replication have been linked to subsequent immune dysregulation, potentially leading to endothelial activation and cytokine storm^19^. Various surrogate markers of viral replication and activation of inflammatory pathways and coagulation cascade have been independently and consistently correlated with worse clinical COVID-19 outcomes^20^. Our findings suggest that proxalutamide, a non-steroidal antiandrogen (NSAA), improves multiple markers of severity in COVID-19, in addition to the reduction in hospitalization rate observed in both sexes.^24,25^

The use of antiandrogens as a promising therapy for COVID-19 is based on the androgen theory on COVID-19. We have previously shown that males, particularly those exhibiting AGA, BPH and or genetic polymorphisms in AR, were more severely affected by COVID-19 ^1,2-5^. Women with hyperandrogenism phenotypes have also been associated with worse prognosis.^7,8^ Finally, we have recently shown that athletes abusing DHT derived AAs may be predisposed to more severe SARS-CoV-2 infections and complications.^12^ Males under androgenic deprivation therapy (ADT) for castration-resistant prostate cancer were linked to lower risk of severe COVID-19, which is unexpected, since ADT may lead to immunosuppression and increased frailty, compared to non-ADT.^9-11^

In an early report of two of our randomized, placebo controlled, double blind clinical trials on Early Anti-androgenic Therapy (EAT) for COVID-19 with two antiandrogens, dutasteride and proxalutamide, we were able to prove the concept that anti-androgenic therapy has a definitive antiviral effect.^17,26^ In the case of proxalutamide, this effect was consistent for both male and females. Proxalutamide antiviral effects may be ascribed both to ACE-2 and TMPRSS-2 down regulation.^21^ The importance of viral load as a surrogate marker has being increasingly recognized, as it directly relates to critical clinical outcomes, such as mechanical ventilation use and death. Unfortunately, because dependence on availability of properly collected, stored and processed rtPCR, the randomization of included patients, originally designed for the primary clinical outcomes, was not able to prevent some imbalances when results were analyzed specifically for viral outcomes.

Here we present a more thorough biochemical analysis without relevant losses to follow-up and attenuation of these previous limitations. In fact, results have been consistent with the biological hypothesis of anti-androgenic agents mechanism of action, and have demonstrated consistency across genders, age, and baselines characteristics.

Proxalutamide therapy was related to improvements in immunologic system, including substantial reduction of neutrophils as early as Day 1, higher increases in total lymphocytes counts, lower N/L ratio, and faster eosinophil increase, as compared to placebo. Strikingly, patients randomized to placebo displayed a progressively increasing N/L ratio, reaching > 3:1 after one week follow up. This demonstrates that proxalutamide may prevent the immunologic progression of COVID-19, since all the changes observed in the proxalutamide group are positive in terms of outcomes. In addition, platelets, allegedly to be directly related with better overall prognosis, raised more rapidly in the proxalutamide group compared to placebo group.

Ferritin levels increase considerably during COVID-19, and has been proposed to be an independent predictor of worse COVID-19 related outcomes.^28,^ Ferritin increased less expressively and decreased more quickly among proxalutamide users compared to placebo, providing an additional sign of proxalutamide anti-inflammatory effects.

We have found that usCRP levels are statistically lower after seven days of proxalutamide therapy when compared to the placebo group, even though patients enrolled to proxalutamide had baseline usCRP levels higher then those enrolled to placebo. CRP is a well-established prognostic factor and despite recent evidence have failed to correlate its levels to circulating IL-6 levels, both markers seems to be independently related to worse clinical outcome in COVID-19.^29,30^ Since androgenic and Il-6 pathway converge, this raise the hypothesis of whether anti-androgenic therapies could induce an indirect anti-inflammatory effect through inhibition of the IL-6 pathway. Remarkably, tocilizumab, a monoclonal antibody directed against IL-6 receptor have recently received Emergency Use Authorization for severe COVID-19 by the FDA, after at least two trials have suggested a small, but statistically significant survival advantage.^30^ Intuitively, proxalutamide could also find reduction in mortality rate. Another inflammatory marker, the 1h-ESR, was also demonstrated to reduce more significantly in proxalutamide group, adding evidence to the anti-inflammatory effects of the drug.

D-dimer is a marker of fibrin degradation and is elevated in diverse pathological thrombogenic processes. Specifically, elevated levels have been associated with higher risk of COVID-19 related death. Major medical societies now recommend anti-coagulation prophylaxis or therapeutics based entirely on D-dimer values.^31^ Patients randomized to proxalutamide exhibited statistically significant lower D-dimer values after seven days of therapy, when compared to placebo. In addition to D-dimer, fibrinogen, another endothelial marker that is directly related to multiple risks, experimented more important reduction in proxalutamide group compared to placebo.

Patients randomized to proxalutamide from both sexes have experienced higher levels of testosterone compared to placebo. In the case of males, this was not predicted to occur due to GnRH agonism action, since the drug does not penetrate CNS markedly. Instead, we hypothesized that proxalutamide could protect Leydig cells from SARS-CoV-2 infections, since these cells have demonstrated to express both ACE-2 and TMPRSS-2. In accordance with this hypothesis, recent work has correlated lower level of testosterone in males to worse prognosis when dosed during COVI-19.^32^ This reinforces the better outcomes observed in this population with proxalutamide. Conversely, the increased testosterone and estradiol levels observed in females is speculated to be related to paracrine actions of AR antagonism in the ovaries and possibly in the adrenal glands.

Although by selection criteria oxygen saturation was normal at baseline, proxalutamide users experimented more important increase than placebo. Whether a mitigation of subclinical endothelial and lung dysfunction that may occur early in COVID-19 played any role in this improvement is unknown.

Our study has some major limitations. First, patients did not receive the same SOC, which was unable to be precisely quantified. We don’t know if proxalutamide would act the same on different SOC backgrounds. However, both groups received the same co-interventions, making confounding bias unlikely. Since our study spanned a period during which different lineages have caused infections in Brazil, it is possible that we have included patients with different viral related factors in distinct enrollment periods. In fact, we have subjectively perceived a progressive increment of baseline usCRP in control group during the study period. Patients included later generally displayed higher usCRP values. However, the randomized nature of the trial would have distributed different lineages evenly, and at worst, this could have lead to a “conservative bias”, since including more severe cases would have shifted results towards null hypothesis. However, we were not able to perform SARS-CoV-2 genotyping and other reasons for usCRP increasing could therefore have happened that could somehow biased our results. Finally, some biochemical parameters have missing data. These were parameters not fully covered by research founding, that have been offered as extra tests randomly. This prevented us to make additional biochemical considerations.

In summary we have found benefits in immunologic, inflammatory and thrombogenic surrogate markers that have been independently linked to clinical outcomes, such as hospital admission, ICU admission, mechanical ventilation and death. Moreover, benefits where equally distributed between male and female patients. Indeed, in our RCTs with proxalutamide for COVID-19 patients demonstrated reduction of hospitalization in both females and males. The reduction of hospitalization rate may have been mediated by the prevention of dysfunctional immunological and inflammatory response that leads to the inflammatory and lung injury stages of COVID-19 In addition, this is on agreement on our view that general hyperandrogenic state may be more important that absolute testosterone levels. Previous work from our group have highlighted that certain AR polymorphisms are related to hype-androgenic states and consequently to more severe COVID-19 disease.

## Conclusion

The substantial improvements observed in immunologic, inflammatory, and thrombotic markers with proxalutamide may support the reduction of hospitalization rate observed in both females and males with COVID-19 using proxalutamide compared to standard of care.

## Data Availability

Dataset is not available.

